# CD4 T cell counts are inversely correlated with anti-cluster A antibodies in antiretroviral therapy-treated PLWH

**DOI:** 10.1101/2025.02.25.25322882

**Authors:** Mehdi Benlarbi, Jonathan Richard, Tommaso Clemente, Catherine Bourassa, William D. Tolbert, Suneetha Gottumukkala, Marc-Messier Peet, Halima Medjahed, Marzena Pazgier, Frank Maldarelli, Antonella Castagna, Madeleine Durand, Andrés Finzi

**Affiliations:** Centre de Recherche du CHUM, Montréal, QC, Canada; Département de Microbiologie, Infectiologie et Immunologie, Université de Montréal, Montreal, QC, Canada; Vita-Salute San Raffaele University, Milan, Italy; Infectious Diseases, IRCCS San Raffaele Scientific Institute, Milan, Italy; Infectious Disease Division, Department of Medicine, Uniformed Services University of the Health Sciences, Bethesda, MD, USA; HIV Dynamics and Replication Program, NCI, NIH, Bethesda, MD 20892, USA; Department of Medicine, Faculty of Medicine, Centre Hospitalier de l’Université de Montréal, Montréal, QC, Canada

**Keywords:** HIV, CD4 T cells, CD4 depletion, soluble gp120, inflammation, immune dysfunction, CD4 binding site antibodies, anti-cluster A antibodies, antibody-dependent cellular cytotoxicity

## Abstract

While antiretroviral therapy efficiently suppresses viral replication, inflammation and immune dysfunction persist in some people living with HIV-1 (PLWH). Soluble gp120 (sgp120) has been detected in PLWH plasma and its presence is linked to immune dysfunction. It was reported that sgp120 binding to CD4 on uninfected bystander CD4^+^ T cells sensitizes them to antibody-dependent cellular-cytotoxicity (ADCC) mediated by non-neutralizing antibodies present in PLWH plasma. Using three independent PLWH cohorts, we observed that non-neutralizing anti-cluster A antibodies are negatively associated with CD4^+^ T cell counts. Anti-CD4BS antibodies blocked the coating of uninfected bystander cells by sgp120, thereby preventing their elimination by ADCC. Supporting a protective role of anti-CD4BS antibodies, PLWH having these antibodies didn’t show a negative association between CD4 T cell counts and anti-cluster A. Our results reveal that anti-cluster A antibodies are associated with immune dysfunction in PLWH and anti-CD4BS antibodies might have a beneficial impact in these individuals.

## Introduction

The hallmark of acquired immunodeficiency syndrome (AIDS) is the progressive depletion of CD4^+^ T cells [1–4]. Antiretroviral therapy (ART) remains the most effective treatment to suppress HIV replication [5–7]. Despite sustainable undetectable viremia, inflammation and immune dysfunction persists in some people living with HIV (PLWH), with ∼10-40% failing to restore their CD4^+^ T cells count [8–11]. Individuals with low CD4^+^ T cells count and/or sustained low CD4:CD8 ratios are at higher risks of developing non-AIDS related malignancies [12–16]. To date, the molecular mechanism by which HIV infection impacts on the immune homeostasis of PLWH with undetectable viremia remains unclear [17, 18].

Viral proteins produced by the persistent HIV reservoir may be one mechanism contributing to these perturbations in long-term ART-treated PLWH [19–22]. One of such antigens is the surface subunit gp120 of the HIV-1 envelope glycoprotein (Env). Env mediates viral entry into host cells and is composed of a trimer of non-covalently linked gp120-gp41 heterodimers [23, 24]. Consequently, gp120 can be spontaneously released from Env trimers expressed at the surface of viral particles or infected cells [25–27]. Soluble gp120 (sgp120) has been detected in blood and tissues from PLWH, even during ART, and its detection has been associated with inflammation and markers of immune dysfunction [28–31].

Prior *in vitro* studies found that the binding of sgp120 to CD4 on immune cells induces the release of pro-inflammatory markers such as IL-6, TNFα, IL-1β and IL-10 [32–34]. Interestingly, PLWH with detectable levels of plasmatic sgp120 had higher levels of circulating inflammatory cytokines compared to sgp120 negative PLWH [28, 30]. Furthermore, highly conserved CD4-induced gp120 epitopes that are normally occluded in the unliganded Env trimer become exposed upon Env ‘’opening’’ or gp120 shedding [35–39]. Binding of sgp120 to CD4 on uninfected bystander CD4^+^ T cells further exposes these vulnerable epitopes, making them a prime target for non-neutralizing antibodies (nnAbs) commonly elicited upon HIV infection [40–43]. Importantly, sgp120 coating of uninfected bystander CD4 T cells sensitizes them to Fc-effector functions, such as antibody-dependent cellular cytotoxicity (ADCC), mediated by plasma from PLWH [32, 41, 43]. Specifically, nnAbs targeting the gp120 inner-domain cluster A region (anti-cluster A Abs) were found to be responsible for most of the ADCC activity of PLWH plasma *in vitro*, provided that their epitopes were exposed [44–47].

We recently reported that small-molecule gp120 inhibitors targeting the CD4 binding-site (CD4BS) can block sgp120 immunomodulatory activities by hindering the gp120-CD4 interaction [32]. Potent anti-CD4BS broadly neutralizing Abs (bNabs) are currently being evaluated in clinical trials for their ability to prevent HIV-1 infection and control viral replication [48, 49]. Here we evaluated in three different cohorts of PLWH whether plasmatic levels of anti-cluster A Abs were associated with CD4^+^ T cell counts and if the presence of anti-CD4BS Abs could modify this association.

## Results

### Uninfected bystander CD4^+^ T cells are eliminated by ADCC responses mediated by plasma from PLWH

To evaluate whether uninfected bystander cells can be eliminated by ADCC responses mediated by plasma from PLWH, we used a previously described co-culture system that allows the distinction between productively infected cells and uninfected bystander cells (Figure 1A) [32, 43, 50]. Briefly, activated primary CD4^+^ T cells were infected with HIV_pNL4.3.ADA.GFP_ virus and incubated with autologous uninfected bystander cells. After two days of co-culture, recognition and ADCC-mediated killing of uninfected bystander cells (eFluor450^+^GFP-) was evaluated using plasma from PLWH or uninfected controls as described previously (Figure 1B-D) [32, 43, 50]. sgp120 released from productively infected cells (eFluor450^-^GFP+) readily binds to uninfected bystander cells (eFluor450^+^GFP-) and becomes efficiently recognized by PLWH plasma (Figure 1B-C). These uninfected bystander cells became susceptible to ADCC responses mediated by PLWH plasma in the presence of autologous peripheral blood mononuclear cell (PBMCs) (Figure 1D).

**Figure 1.**
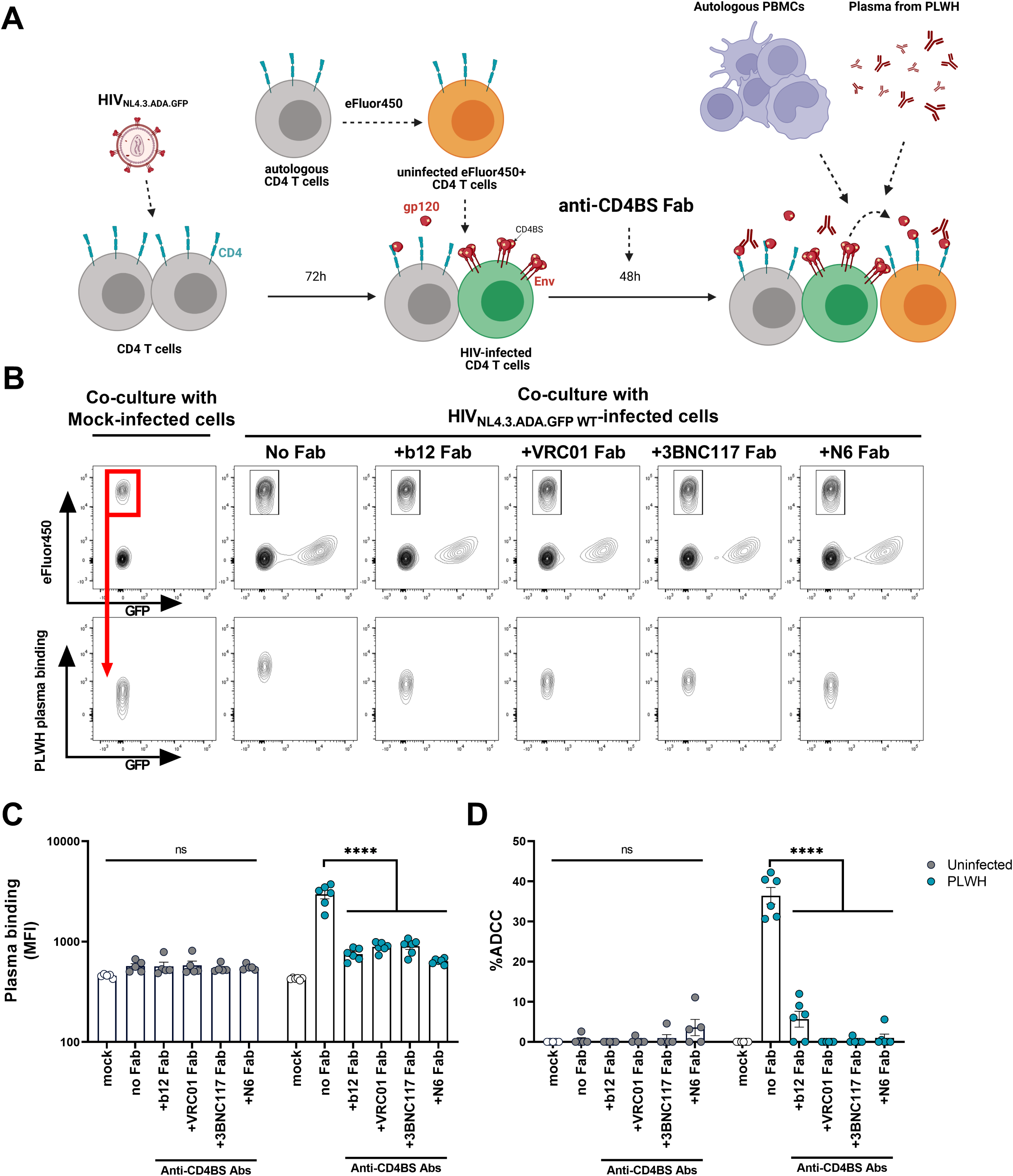
Uninfected bystander CD4+ T cells are eliminated by ADCC responses mediated by plasma from PLWH. (A) Activated primary CD4+ T cells were infected with GFP reporter HIV_NL4.3.ADA.GFP WT_ virus. Three days post infection, autologous uninfected CD4+ T cells were stained with the eFluor450 dye and co-cultured with mock-infected or HIV-1-infected cells (eFluor450^-^ GFP+ cells) for 2 days in the presence or absence of anti-CD4BS Fab fragments (20 µg/mL) (B–D). (C) Recognition and (D) ADCC-mediated elimination of uninfected bystander cells, designated eFluor450^+^GFP-cells, by plasma from 5 uninfected or 6 PLWH individuals were evaluated by flow cytometry. Shown in (B) are the gating strategy and representative staining obtained with PLWH plasma. The graphs shown represent (C) the median fluorescence intensity (MFI) and (D) the percentage of ADCC obtained with different PLWH plasmas. Error bars indicate means ± standard error of the mean (SEM). Statistical significance between no Fab and with Fab conditions was tested using a two-way ANOVA with a Holm-Sidak’s post-test (*p < 0.05, **p < 0.01, ***p < 0.001, ****p <0.0001).

We then evaluated whether anti-CD4BS antibodies, some of which are currently used in clinical trials [48], could block the sgp120-mediated killing of uninfected bystander CD4^+^ T cells. Fab portions of b12 [51], VRC01 [52], 3BNC117 [53] and N6 [54] were added to the co-culture system. The presence of anti-CD4BS Fabs abrogated sgp120-coating of uninfected bystander cells and subsequently prevented their elimination by ADCC responses mediated by PLWH plasma (Figure 1C-D). As expected, uninfected bystander cells were not recognized, nor killed by plasma from uninfected controls (Figure 1C-D).

### Plasma from PLWH with anti-CD4BS Abs protect uninfected bystander CD4^+^ T cells from ADCC responses

It is well established that some, but not all PLWH have anti-CD4BS in their plasma [55, 56]. The differential elicitation of these antibodies remains poorly understood [57]. Since CD4BS Fab fragments were able to block the coating of uninfected bystander cells by sgp120 and therefore prevent their elimination by ADCC mediated by plasma from PLWH (Figure 1), we asked whether endogenously elicited CD4BS Abs could modulate this response. First, we measured by ELISA the levels of anti-CD4BS Abs using the RSC3 probe, known to specifically bind to CD4BS Abs [52, 58, 59]. We used plasma from 386 participants from the Canadian HIV and Aging Cohort Study (CHACS) (Table S1). In this cohort of long-term ART treated PLWH, we found that 66/386 (17.1%) participants had detectable levels of anti-CD4BS Abs (Table S2).

We then evaluated the capacity of the plasma of 37 participants with or without anti-CD4BS Abs to prevent sgp120-coating and ADCC-mediated killing of uninfected bystander CD4^+^ T cells using a modified flow-cytometry based co-culture assay (Figure 2A) [32]. In this assay, recombinant monomeric sgp120 was pre-incubated with plasma containing or not anti-CD4BS Abs. sgp120 binding to the surface of uninfected bystander cells (eFluor450^+^CFSE-) was measured by flow cytometry (Figure 2A-B). In PLWH without anti-CD4BS Abs, sgp120-immunocomplexes readily bound to these cells, which subsequently became susceptible to ADCC responses mediated by autologous PBMCs (Figure 2B-C). Strikingly, we observed that the presence of endogenously elicited anti-CD4BS Abs reduced sgp120-coating and ADCC-mediated killing of uninfected bystander cells (Figure 2B-C). To rule out the possibility that these differences were due to lower levels of anti-cluster A Abs in these participants, we measured the levels of these Abs using an engineered stabilized gp120 inner domain 2 protein (ID2) probe, which exposes only the cluster A region [60, 61]. We found no differences in the quantity of nnAbs in both groups (Figure S1). These results suggest that the presence of anti-CD4BS Abs in PLWH plasma may prevent CD4^+^ T cells depletion mediated by sgp120 and nnAbs.

**Figure 2.**
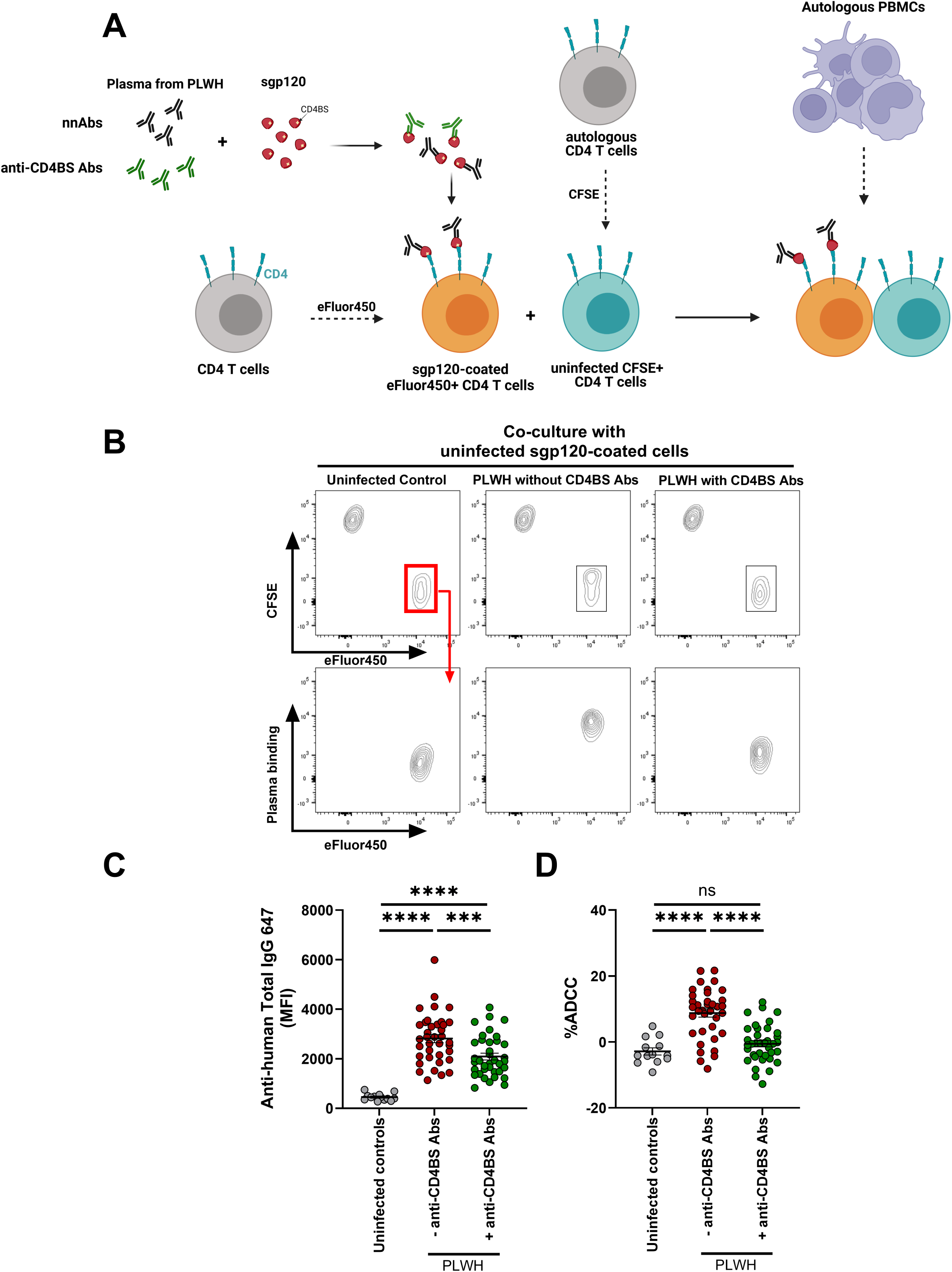
Plasma from PLWH with anti-CD4BS Abs protect uninfected bystander CD4+ T cells from ADCC responses. (A) Activated primary CD4+ T cells stained with the eFluor450 dye and then coated with sgp120-Abs complexes. These coated cells were subsequently co-cultured with uncoated (eFluor450-CFSE+ cells) at a 1:1 ratio. (B–D). (C) Recognition and (D) ADCC-mediated elimination of coated cells, designated eFluor450^+^CFSE-cells, by plasma from 13 uninfected or 37 PLWH individuals with or without anti-CD4BS Abs were evaluated by flow cytometry. Shown in (B) are the gating strategy and representative staining obtained with PLWH plasma. The graphs shown represent (C) the median fluorescence intensity (MFI) and (D) the percentage of ADCC obtained with different PLWH plasmas. Error bars indicate means ± standard error of the mean (SEM). Statistical significance between conditions was tested using an ordinary one-way ANOVA with a Holm-Sidak’s post-test (ns, not significant, ***p < 0.001, ****p <0.0001). See also Figure S1.

### Anti-cluster A antibodies are negatively associated with CD4 T cell counts in PLWH and anti-CD4BS antibodies abrogate this association

To evaluate whether the level of anti-cluster A Abs is associated with CD4^+^ T cell counts *in vivo*, we measured their levels in three distinct cohorts of PLWH (CHACS, PRESTIGIO and NIH). CHACS is a Canadian prospective study enrolling participants who are aged 40 years or older or have lived with HIV for 15 years or more (Table S1) [62]. The PRESTIGIO Registry is an ongoing Italian prospective cohort enrolling people with 4-class drug-resistant HIV (4DR-PLWH), with 25% of tested participants having detectable viremia despite ART usage (Table S1) [63]. Lastly, the NIH cohort (AVBIO2, protocol 08-I-0221) is comprised of PLWH enrolled in a natural history study of the effects of ART in adult PLWH at the NIH Clinical Center in Maryland (US) (Table S1). Overall, participants had a long history of HIV infection and were under long-term ART treatment, with most of them being virally suppressed, as summarized in Table S1. PRESTIGIO participants had lower CD4:CD8 ratios and Nadir CD4. NIH participants had a higher proportion of black participants. No major differences in age and biological sex among the three cohorts were noted.

In the three cohorts we observed a negative association between anti-cluster A Abs levels and CD4 T cell counts, albeit this association was only significant for the CHACS cohort (Figure 3A, Table S3, S4). We then evaluated the presence of anti-CD4BS Abs in these individuals and stratified the analysis based on the absence or presence of these Abs. There were no differences in terms of demographics or immune parameters between those with or without anti-CD4BS Abs (Table S2). Remarkably, the negative association between anti-cluster A Abs and CD4 T cell counts became more pronounced and significant in individuals that didn’t have anti-CD4BS Abs, pointing to a potential beneficial role of anti-CD4BS Abs in these individuals (Figure 3B, Table S3, S4). Further supporting this hypothesis, no association was observed between anti-cluster A Abs and CD4 T cell counts in PLWH having anti-CD4BS Abs (Figure 3C, Table S3, S4).

**Figure 3.**
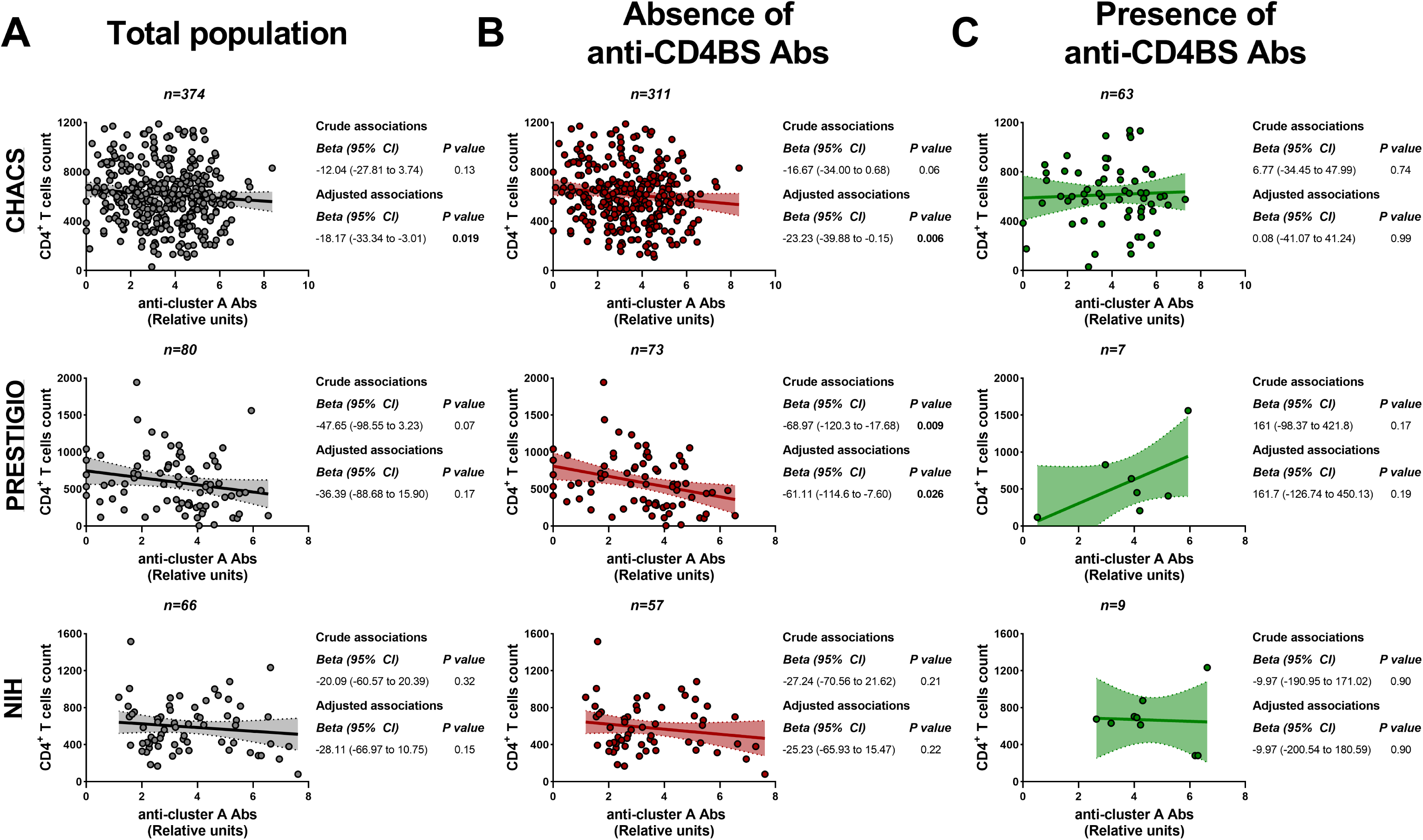
Anti-cluster A antibodies are negatively associated with CD4 T cell counts in PLWH and anti-CD4BS antibodies abrogate this association. Correlations between CD4^+^ T-cell counts and anti–cluster A antibody levels are shown for the three cohorts tested (CHACS; upper row, PRESTIGIO; middle row and NIH; bottom row). Correlations are shown for (A) the total study population or stratified by (B) the absence or (C) presence of anti-CD4BS. Levels of anti–cluster A antibodies were log_2_ transformed. The bold line represents a simple linear regression with the 95% confidence bands shown in the dotted line, and shaded in grey (total population), red (absence of anti-CD4BS Abs) or green (presence of anti-CD4BS Abs). Univariable and multivariable linear regressions were performed, with the beta parameters representing the mean predicted change in absolute CD4^+^ T cell counts for each 1-log_2_ increase in titers of anti–cluster A antibodies. Multivariable models are adjusted for age, sex, duration of antiretroviral therapy and nadir CD4 counts. See also Table S3 and S4.

We further evaluated the strength of these associations by combining the three cohorts together. To do this, we used generalized least squares linear regression model with random effects (considering clustering within cohorts) and adjusted for potential confounding factors such as age, sex, ART duration and nadir CD4. We formally assessed how this association was modified by anti-CD4BS Abs by adding an interaction term to the model between the absence or presence of anti-CD4BS Abs and anti-cluster A Abs (Table 1, Table S5). Within the total study population, each 1-log_2_ increase in levels of anti-cluster A Abs was associated with a mean predicted decrease in CD4^+^ T cell count of −23.85 (95% confidence interval [CI], −37.75 ×10^6^/mL to −9.95 ×10^6^/mL; *p* = 0.001). The magnitude of this association was more pronounced in the subgroup without anti-CD4BS Abs, with a mean predicted decline in CD4^+^ T cell count of −31.91 (95% [CI], −46.97 ×10^6^/mL to −16.84 ×10^6^/mL; *p* < 0.001). This negative association was lost in participants with anti-CD4BS Abs with a mean predicted change of 13.10 (95% [CI], −26.35 ×10^6^/mL to 52.55 ×10^6^/mL; *p* = 0.51). This difference in effects between the groups was statistically significant (p value for interaction 0.03) (Table 1). Altogether, we observed that the presence of anti-CD4BS in PLWH plasma modified the association between anti-cluster A Abs and CD4^+^ T cells count, likely through preventing sgp120-coating on bystander CD4^+^ T cells.

**Table 1.** Pooled adjusted associations between anti-cluster A Abs and CD4 count, overall and stratified by the absence or presence of anti-CD4BS Abs.

|  |  |  |  |  |
| --- | --- | --- | --- | --- |
|  | n | Beta*, 95% CI | P-value | p-value for interaction |
| Overall** | 520 | -23.85 [-37.75 to -9.95] | 0.001 | NA |
| Absence of anti-CD4BS Abs | 441 | -31.91 [-46.97 to -16.84] | <0.001 | 0.03 |
| Presence of anti-CD4BS Abs | 79 | 13.10 [-26.35 to 52.55] | 0.51 |  |

\*beta coefficients represent the predicted change in CD4 count for each increase of 1-log<sub>2</sub> in levels of anti-cluster A antibodies
\*\*12 participants did not have information on CD4 counts and were excluded from the analysis
All models are adjusted for age, sex, duration of antiretroviral therapy and nadir CD4
Abbreviations: CI; confidence intervals, anti-CD4BS Abs; anti-CD4 binding site antibodies
Also refer to Table S5 for crude associations.

## Discussion

Persistent inflammation, immune activation and incomplete CD4^+^ T cell recovery remain a critical challenge to achieving successful aging among virally suppressed PLWH [17, 64]. Such immune perturbations have been associated with higher morbidity and mortality [13–15]. These conditions can range from cardiovascular disease [65, 66], renal disease [67], liver disease [68], bone disease [69], neurological conditions [70] to cancers [71], all of which typically appear sooner to PLWH than to the general population [16, 17]. Here we report that anti-cluster A Abs are negatively associated with CD4^+^ T cell counts and that the presence of anti-CD4BS Abs abrogates this negative association.

Previous *in vitro* experiments have shown that shed gp120 can sensitize uninfected bystander CD4^+^ T cells to elimination by ADCC mediated by nnAbs, particularly those targeting the gp120 cluster A region (anti-cluster A Abs) [41, 43]. Interestingly, administration of the gold standard anti-cluster A mAb A32 in HIV_JRSCF_-infected NSG-15 PBL humanized-mice led to the depletion of CD4^+^ T cells in blood and several tissues, suggesting that ADCC-mediated killing of uninfected bystander cells could also happen *in vivo* [72]. Here we observed that despite long-term ART treatment, 98.3% of participants tested had detectable levels of anti-cluster A Abs. The levels of these antibodies were associated with a significant decline in the CD4 count. Importantly, we did not observe this association in participants with anti-CD4BS Abs, raising the possibility that blocking the gp120-CD4 interaction might provide a immunological benefit beyond blocking the viral entry.

Currently, several potent bNabs are being evaluated in large scale clinical trials for treatment and prevention of HIV infection through passive immunization [48, 49]. In this study, we observed that anti-CD4BS mAbs b12, VRC01, 3BNC117 and N6 protected uninfected bystander CD4^+^ T cells from ADCC-mediated killing. Interestingly, administration of anti-CD4BS bNabs during ART initiation or interruption in PLWH has been reported to induce a ‘’vaccinal effect’’ [73–76]. This effect is described as a durable enhancement of HIV-specific CD4 and CD8 T cells responses, with some studies also describing an improvement in humoral responses [77]. Whether this host enhancement of adaptive responses is associated with the protection of CD4^+^ T cells by preventing their sgp120-mediated elimination remains to be determined.

It is well established that ART does not preclude transcription and translation of viral products within cells harboring intact or defective HIV proviruses [19, 22, 78–80]. While most infected cells persisting under ART harbor defective ‘’replication incompetent’’ proviruses, their capacity to produce viral proteins, trigger immune activation and maintain HIV-specific T cells responses have been observed in several studies [20, 21, 81–84]. It has been shown in some cohorts that anti-HIV antibodies correlate positively with the size of the HIV reservoir [28, 85, 86]. Strategies aimed at characterizing the persistent viral reservoir, particularly those endowed with the capacity to express Env, are needed to better understand the source of sgp120 and the persistence of anti-cluster A Abs and anti-CD4BS Abs despite undetectable viremia.

Altogether, preventing nnAbs-mediated killing of uninfected CD4^+^ T cells by targeting sgp120 may represent a promising approach to alleviate symptoms of inflammation and immune dysfunction in PLWH under ART.

## Limitations of the study

The anti-CD4BS mAbs used in this study are broadly neutralizing antibodies that only a minority of PLWH elicit. PLWH tested in this study were mostly virally suppressed, whether anti-CD4BS Abs elicited during acute, early or chronic HIV infection block sgp120 immunomodulatory activities remains to be determined. Further, only a small proportion of PLWH tested in this study had detectable levels of anti-CD4BS Abs, validation with larger cohorts will be helpful to establish the benefits of these Abs.

## Data Availability

All data produced in the present work are contained in the manuscript

## Acknowledgments

The authors thank the CRCHUM BSL3 and Flow Cytometry Platforms for technical assistance. Figure 1A and Figure 2A were prepared using illustrations from BioRender. The following reagents were obtained through the National Institutes of Health HIV Reagent Program, Division of AIDS, National Institute of Allergy and Infectious Disease: Anti-Human Immunodeficiency Virus (HIV)-1 gp120 Monoclonal Antibody (IgG1 b12), contributed by Dr. Dr. Dennis Burton and Dr. Carlos Barbas; Human Immunodeficiency Virus 1 (HIV-1) VRC01 Monoclonal Antibody Heavy and Light Chain Expression Vectors contributed by John Mascola; N6 monoclonal Antibody Heavy and Light Chain Expression Vectors contributed by Drs Jinghe Huang and Mark Connors; and resurfaced stabilized core 3 protein, recombinant from HEK293 cells contributed by Drs Zhi-Yong Yang, Peter Kwong, Gary Nabel, and John Mascola. We also thank Michel Nussenzweig (The Rockefeller University) for providing the plasmids for 3BNC117 and James Robinson for providing the plasmids for A32 and C11. This study was supported by a CIHR Team grant #197728, a Canada Foundation for Innovation (CFI) grant #41027 to A.F and by the National Institutes of Health to A.F. (R01AI148379, R01AI176531), A.F., and M.P. (R01AI174908, R01AI150322), A.F., and M.P. (R01AI186809), M.P. (P01AI162242). Support for this work was also provided by UM1AI164562 (ERASE) to A.F. M.D. received a clinician-researcher salary award from Fonds de Recherche du Québec-Santé. The CHACS cohort is supported by a CIHR (grant HAL-157985) and CIHR’s HIV Clinical Trial Network (Grant CTNPT-052.). F.M. received support from intramural NIH funding (ZIA BC 011466). M.B is recipient of a doctoral CIHR fellowship (FBD-193357). The funders had no role in study design, data collection and analysis, decision to publish, or preparation of the manuscript.

## Author contributions

Conceptualization, A.F., M.D., A.C., F.M., and M.B.; Methodology, M.B., J.R., C.B., H.M. and A.F.; Investigation, M.B., J.R., T.C., C.B., M-M.P., H.M., and F.M.; Formal analysis, M.B., and M.D.; Visualization: M.B., and M.D.; Writing – Original Draft, M.B., and A.F.; Writing – Review & Editing, M.B., J.R., T.C., C.B., W.D.T., M-M.P., H.M., M.P., F.M., A.C., M.D., and A.F.; Funding Acquisition, A.F., M.D., A.C., and F.M.; Resources, M.B., J.R., T.C., S.K., W.D.T., M-M.P., M.P., F.M., A.C., M.D., and A.F.; Supervision, A.F., M.D., A.C., F.M., and M.B.

## STAR METHODS

### RESOURCE AVAILABILITY

#### Lead contact

Further information and requests for resources and reagents should be directed to and will be fulfilled by the lead contact, Andrés Finzi.

#### Materials availability

All unique reagents generated in this study are available from the lead contact with a completed Materials Transfer Agreement.

#### Data and code availability

- All data reported in this paper will be shared by the lead contact upon request
- This paper does not report original code
- Any additional information required to reanalyze the data reported in this paper is available from the lead contact upon request

### EXPERIMENTAL MODELS AND SUBJET DETAILS

#### Ethics statement

Written informed consent was obtained from all study participants. Research adhered to the ethical guidelines of CRCHUM and was reviewed and approved by the CRCHUM institutional review board (ethics committee, approval number CE 16.164 - CA). The PRESTIGIO Registry was approved on December 14, 2017 (Clinical trial, NCT 04098315). The NIH cohort (AVBIO2, protocol 08-I-0221) was approved by the National Institutes of Health IRB#10 (IRB00011862), FWA00005897. Research adhered to the standards indicated by the Declaration of Helsinki.

#### Cell lines and primary cells

HEK293T human embryonic kidney cells (obtained from ATCC) were maintained at 37⁰C under 5% CO_2_ in Dulbecco’s Modified Eagle Medium (DMEM) (Wisent, St. Bruno, QC, Canada), supplemented with 5% fetal bovine serum (FBS) (VWR, Radnor, PA, USA) and 100 U/mL penicillin/streptomycin (Wisent). FreeStyle 293F cells (Thermo Fisher Scientific) were grown in FreeStyle 293F medium (Thermo Fisher Scientific) to a density of 1 × 10^6^ cells/mL at 37°C with 8% CO2 with regular agitation (150 rpm). Primary human PBMCs and CD4+ T cells were isolated, activated and cultured as previously described [36]. Briefly, PBMCs from uninfected controls; 1 male (47 years of age) and 1 female (44 years of age) were obtained by leukapheresis and Ficoll-Paque density gradient isolation were cryopreserved in liquid nitrogen until further use. CD4^+^ T lymphocytes were purified from resting PBMCs by negative selection using immunomagnetic beads per the manufacturer’s instructions (StemCell Technologies, Vancouver, BC) and were activated with phytohemagglutinin-L (10 mg/mL) for 48 h and then maintained in Roswell Park Memorial Institute (RPMI) 1640 complete medium supplemented with rIL-2 (100 U/mL).

### METHOD DETAILS

#### Plasmids and proviral constructs

The vesicular stomatitis virus G (VSV-G)-encoding plasmid has been previously described [87]. The plasmid pSVIIIenv expressing the full-length HIV-1_YU2_ Env WT and the Tat-expressing plasmid (pLTR-Tat) were as previously reported [27]. The infectious molecular clones (IMC) HIV-_1NL4.3.ADA.GFP_ WT was previously described [36, 88].

#### Viral production and infections

VSV-G-pseudotyped HIV-1 viruses were produced by co-transfection of 293T cells with an HIV-1 proviral construct and a VSV-G-encoding vector using the PEI method. Two days post-transfection, cell supernatants were harvested, clarified by low-speed centrifugation (300xg for 5 min), and concentrated by ultracentrifugation at 4⁰C (100,605xg for 1 h) over a 20% sucrose cushion. Pellets were resuspended in fresh complete RPMI-1640 media, and aliquots were stored at −80⁰C until use. Virus was then used to infect activated primary CD4+ T cells from uninfected HIV-1 negative donors by spin infection at 800xg for 1 h in 96-well plates at 25⁰C. Viral preparations were titrated directly on primary CD4+ T cells to achieve around 10-15% GFP+ cells.

#### Antibodies and PLWH plasma

Goat anti-human IgG (H+L) antibodies pre-coupled to Alexa Fluor 647 (Invitrogen, Rockford, IL, USA) was used as secondary Abs in flow cytometry experiments. To measure plasma binding at the surface of primary CD4^+^ T cells in presence of anti-CD4BS Fabs, goat anti-human IgG Fc pre-coupled to Alexa Fluor 647 (Invitrogen, Rockford, IL, USA) was used. For ELISA, goat anti-human IgG pre-coupled to HRP (Invitrogen, Rockford, IL, USA) was used. Plasma from CHACS, PRESTIGIO and NIH were collected, heat-inactivated and conserved at −80⁰C until use for flow cytometry and ELISA.

#### Anti-CD4BS Fab preparation

Anti-CD4BS Fabs were prepared by papain digest of anti-CD4BS IgG. Briefly, purified IgG was buffer exchanged into Fab digest buffer, 10 mM sodium phosphate and 5 mM cysteine pH 7.2. IgG was then added to immobilized papain (Thermo Fisher) and incubated at 37° C for 3-4 hours. Papain was removed by centrifugation and the supernatant filtered through a 0.2-micron syringe filter. Fab was separated from Fc and undigested IgG by passage over a HiTrap protein A HP column (cytiva) equilibrated in phosphate buffered saline (PBS) pH 7.2. Flow through fractions were combined, concentrated and the buffer exchanged into PBS. Fab purity was monitored by SDS-PAGE.

#### Co-culture assay with infected cells

The co-culture of HIV_pNL4.3.ADA.GFP-WT_ infected cells with autologous uninfected CD4+ T cells was performed as previously described [32, 43, 50]. Briefly, activated primary CD4+ T cells were infected with VSV-G pseudotyped HIV_pNL4.3.ADA.GFP-WT_ virus. Three days post-infection, autologous uninfected CD4+ T cells were stained with the eFluor450 dye (1:1000 dilution, eBiosciences) for 15 min at room temperature and washed twice with complete RPMI-1640 media before being co-cultured with infected cells at a ratio of 1 uninfected cell to 4 infected cells at 37⁰C, 5% CO_2_, in the presence or absence of anti-CD4BS Abs Fab fragments (20µg/mL) or equivalent volume of PBS. After two days of co-culture, cells were stained with PLWH plasma from 6 individuals (1:1000 dilution) within the CHACS cohort followed with appropriate secondary Abs. The uninfected bystander cells were designated as the eFluor450^+^GFP-cells. To measure ADCC responses against uninfected bystander cells, cells from the co-culture were stained with a viability dye (AquaVivid; Thermo Fisher Scientific) while autologous PBMCs effectors cells were stained with the eFluor670 cell marker (eBioscience). PBMCs were added to target cells at an effector: target ratio of 10:1 in 96-well V-bottom plates (Corning, Corning, NY). PLWH plasma (1:1000 final dilution) were added to appropriate wells and cells were incubated for 15 min at room temperature. The plates were subsequently centrifuged for 1 min at 300xg, and incubated at 37⁰C, 5% CO_2_ for 5 to 6 h before being fixed with a PBS-formaldehyde solution (2% formaldehyde final concentration) containing a constant number of flow cytometry particles (5 x 10^4^ /mL) (AccuCount Blank Particles, 5.3 µm; Spherotech, Lake Forest, IL, USA). As previously reported [43] these flow cytometry particles are designed to be used as reference particle and were used to calculate the absolute number of viable uninfected bystander cells after incubation with autologous PBMCs and PLWH plasma. The percentage of ADCC responses directed against the uninfected bystander cell population (eFluor450^+^eFluor670^-^GFP-viable cells) was calculated with the following formula: ((number of uninfected bystander cells in the Targets + PBMCs condition) - (number of uninfected bystander cells in the Targets + PBMCs + plasma condition))/(number of uninfected bystander cells in the Targets alone condition). Samples were acquired on an LSRII cytometer (BD Biosciences), and data analysis was performed using FlowJo v10.8.1 (Tree Star, Ashland, OR, USA).

#### Protein production and purification

Production and purification of monomeric soluble HIV-1_YU2_ gp120 was described elsewhere [28, 32, 89]. Briefly, recombinant HIV-1_YU2_ gp120 was produced using a plasmid (pcDNA3.1) encoding the codon-optimized full-length HIV-1_YU2_ gp120 containing a C-terminal hexa-histidine tag [26, 90]. FreeStyle 293F cells were transfected with the gp120 expressor using ExpiFectamine 293 transfection reagent, as directed by the manufacturer (Thermo Fisher Scientific). One week later, cells were pelleted, and supernatants were filtered using a 0.22-μm-pore-size filter (Thermo Fisher Scientific). Recombinant gp120 was purified by nickel affinity columns, as directed by the manufacturer (Thermo Fisher Scientific). Monomeric gp120 was subsequently purified by fast protein liquid chromatography (FPLC), as previously reported [89]. The purification by FPLC was performed using an ÄKTAprime Plus FPLC with a HiLoad 16/60 Superdex 200 PG (GE Healthcare, Chicago, IL, USA). The gp120 preparations were dialyzed against phosphate-buffered saline (PBS) and stored in aliquots at −80°C until further use. To assess purity, recombinant proteins were loaded on non-reducing SDS-PAGE polyacrylamide gels and stained with Coomassie blue.

#### Co-culture assay with sgp120-Abs coated CD4^+^ T cells

To prepare sgp120-Abs complexes, 200 ng/mL of monomeric recombinant HIV_YU2_ gp120 was mixed at a 1:1 ratio with plasma from PLWH or uninfected controls from the CHACS cohort diluted at 1:500 and subsequently incubated for 30 min at room temperature. These sgp120-Abs complexes were then added to pre-stained eFluor450 activated primary CD4+ T cells resuspended at 1 x 10^6^ cells/mL for 30 min at 37⁰C, 5% CO_2_. In parallel, autologous uncoated CD4^+^ T cells were stained with the CFSE dye (1:1000 dilution, Invitrogen) for 15 min at room temperature and washed twice with complete RPMI-1640 media. Uncoated cells (CFSE+eFluor450-) were mixed with coated cells (CFSE-eFluor450+) at a 1:1 ratio. These cells were subsequently stained with goat anti-human IgG Alexa Fluor 647. To measure ADCC responses, autologous PBMCs effectors cells were stained with the eFluor670 cell marker (eBioscience). PBMCs were added to target cells at an effector: target ratio of 10:1 in 96-well V-bottom plates (Corning, Corning, NY) and subsequently centrifuged for 1 min at 300xg, and incubated at 37⁰C, 5% CO_2_ for 5 to 6 h before being fixed with a PBS-formaldehyde solution (2% formaldehyde final concentration). The percentage of ADCC responses directed against the coated cell population (eFluor450^+^CFSE-) was calculated with the following formula: ((percentage of eFluor450^+^CFSE-cells in the Target + PBMCs condition) - (percentage of eFluor450^+^CFSE-cells in the Targets + PBMCs + plasma condition))/(percentage of eFluor450+CFSE-cells in the Targets alone condition). Samples were acquired on an LSRII cytometer (BD Biosciences), and data analysis was performed using FlowJo v10.8.1 (Tree Star, Ashland, OR, USA). PLWH plasma used for this experiment were randomly selected among participants with (n=66) or without (n=320) anti-CD4BS Abs within the CHACS cohort.

#### Enzyme-linked immunosorbent Assay

The ELISAs used in this study were described elsewhere [28]. For measurement of anti-cluster A antibodies, wells were coated with stabilized gp120 inner domain ID2 [60] (0.1 µg/mL in PBS), in parallel with BSA (0.1 µg/mL in PBS). After blocking, the cluster A specific A32 mAb (1 µg/mL) or diluted plasma 1:1000 from PLWH or uninfected individuals were added to the well and detection of plasma antibodies was performed using HRP-conjugated goat-anti-human IgG (Invitrogen) at a dilution of 1:3000. HRP enzyme activity was determined after the addition of a 1:1 mix of Western Lightning oxidizing and luminol reagents (Perkin Elmer Life Sciences, Waltham, MA, USA). Light emission was measured with an LB942 Tri-Star luminometer (Berthold Technologies, Bad Wildbad, Germany). Signal obtained with BSA was subtracted for each plasma and were then normalized to the signal obtained with A32 mAb present in each plate. The positivity threshold was established using the following formula: mean of 20 plasmas from uninfected donors + (3 standard deviation of the mean of the 20 plasmas from uninfected donors). The positivity thresholds were calculated with uninfected donors from each respective cohort (CHACS, PRESTIGIO and NIH).

For measurement of CD4-binding site (CD4Bs) antibodies, wells were coated with resurfaced stabilized core 3 (RSC3) [52] (0.1 µg/mL in PBS). We also coated wells in parallel with BSA (0.1 µg/mL in PBS). After blocking, 2G12 mAb (1 µg/mL), VRC01 mAb (1 µg/mL) or diluted plasma 1:1000 from PLWH or uninfected individuals were added to the well and detection of plasma antibodies was performed using HRP-conjugated goat-anti-human IgG (Invitrogen) at a dilution of 1:3000. Signal was measured as described above and signal obtained with BSA was subtracted for each plasma and were then normalized to the signal obtained with 2G12 mAb present in each plate. The positivity threshold was established using the following formula: mean of 20 plasmas from uninfected donors + (3 standard deviation of the mean of the 20 plasmas from uninfected donors. The positivity thresholds were calculated with uninfected donors from each respective cohort (CHACS, PRESTIGIO and NIH).

### QUANTIFICATION AND STATISTICAL ANALYSIS

Every variable was tested for statistical normality, and this information was used to apply the appropriate (parametric or nonparametric) statistical test and descriptive statistics. To assess the association between anti-cluster A antibody levels and CD4^+^ T cell counts within each cohort separately, we used uni- and multi-variable linear regression models, with log_2_-transformed levels of anti-cluster A antibodies as the independent variable and CD4 counts as the dependent variable. The beta coefficients and 95% confidence interval reported represent the model-predicted change for each 1-log increase in anti-cluster A antibodies levels. Potential confounders were identified using a priori clinical knowledge and included age, sex, nadir CD4 and duration of antiretroviral therapy (in years) and were added to multivariable models. When small groups made parsimony necessary, potential confounders were entered sequentially into the models and kept if they modified the point estimate for the association of interest by 10% or more. To assess the presence of effect modification by the absence or presence of anti-CD4BS Abs on the association of interest, an interaction term was added to the models (anti-cluster A Abs*anti-CD4BS Abs). Missing data for the potential confounding factors (age and sex had no missing values, 83 participants had missing nadir CD4 values and 26 had missing ART duration) were handled using multiple imputation with chained equation with 50 imputations for each missing data point. No data was imputed for the outcome or exposures of interest. To assess the association between anti-cluster A Abs and CD4 counts in the three cohorts combined, we used multivariable generalized least squared models, taking into account the clustering within cohorts. Adjustment for confounding factors, handling of missing data and testing for interaction with anti-CD4BS Abs were done as described above.

For all analyses, p values < 0.05 were considered significant, with no adjustment done for multiple testing. Significance values in figures are indicated as *p < 0.05, **p < 0.01, ***p < 0.001, ****p < 0.0001. Analyses were done using GraphPad Prism version 10.4.0 (GraphPad, San Diego, CA, USA) and Stata v 17 (StataCorp. 2021. *Stata Statistical Software: Release 17*. College Station, TX: StataCorp LLC.)

## Supplemental information

**Supplemental Figure 1.**
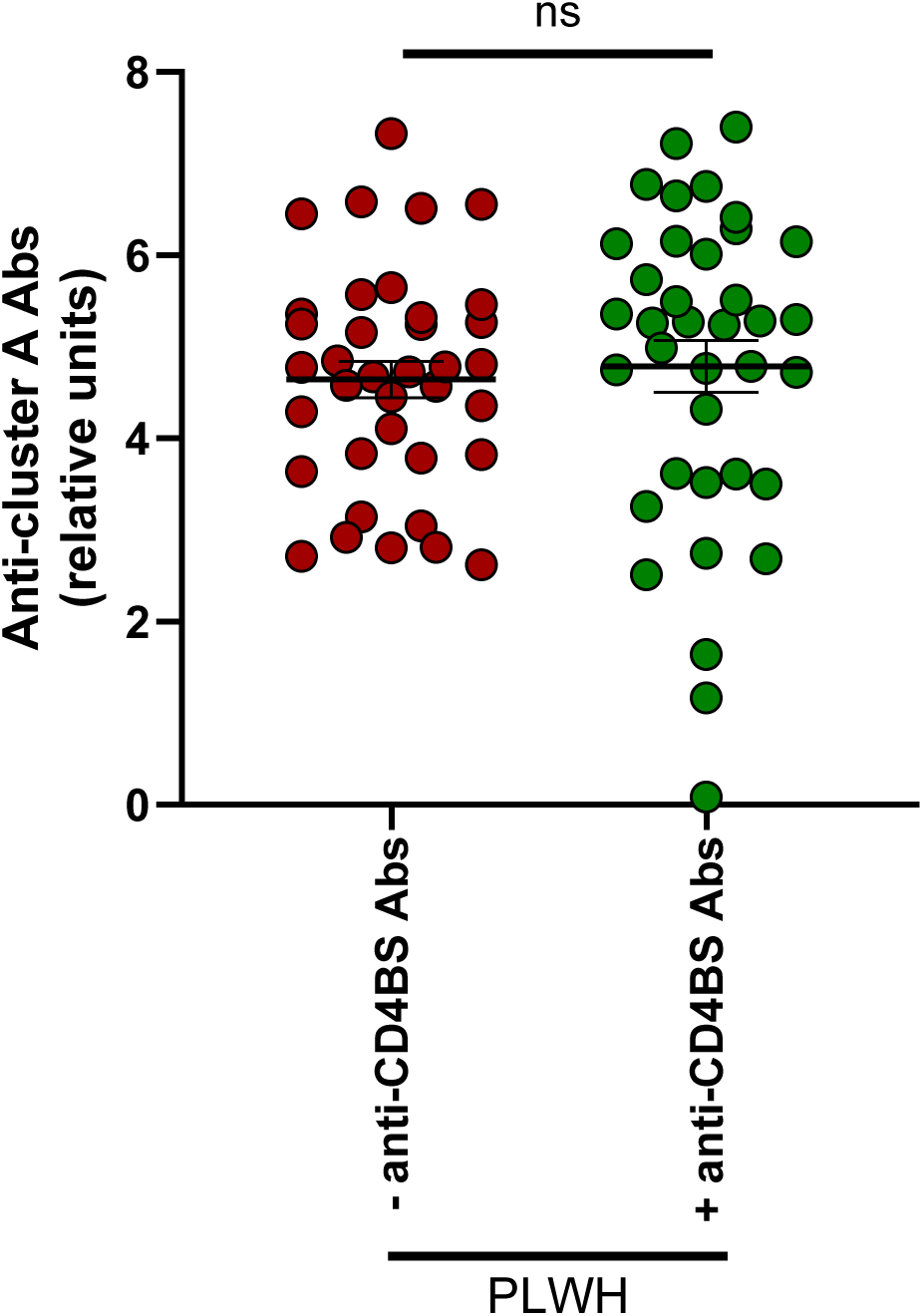
Levels of anti-cluster A Abs in participants with or without anti-CD4BS Abs. Levels of anti-cluster A Abs for 37 PLWH with or without anti-CD4BS Abs within the CHACS cohort were measured by ELISA using the gp120 ID2 probe as described previously. Error bars indicate means ± standard error of the mean (SEM). Statistical significance between the two groups was tested using an unpaired t-test (ns, not significant). Related to Figure 2.

**Supplemental Table 1.**
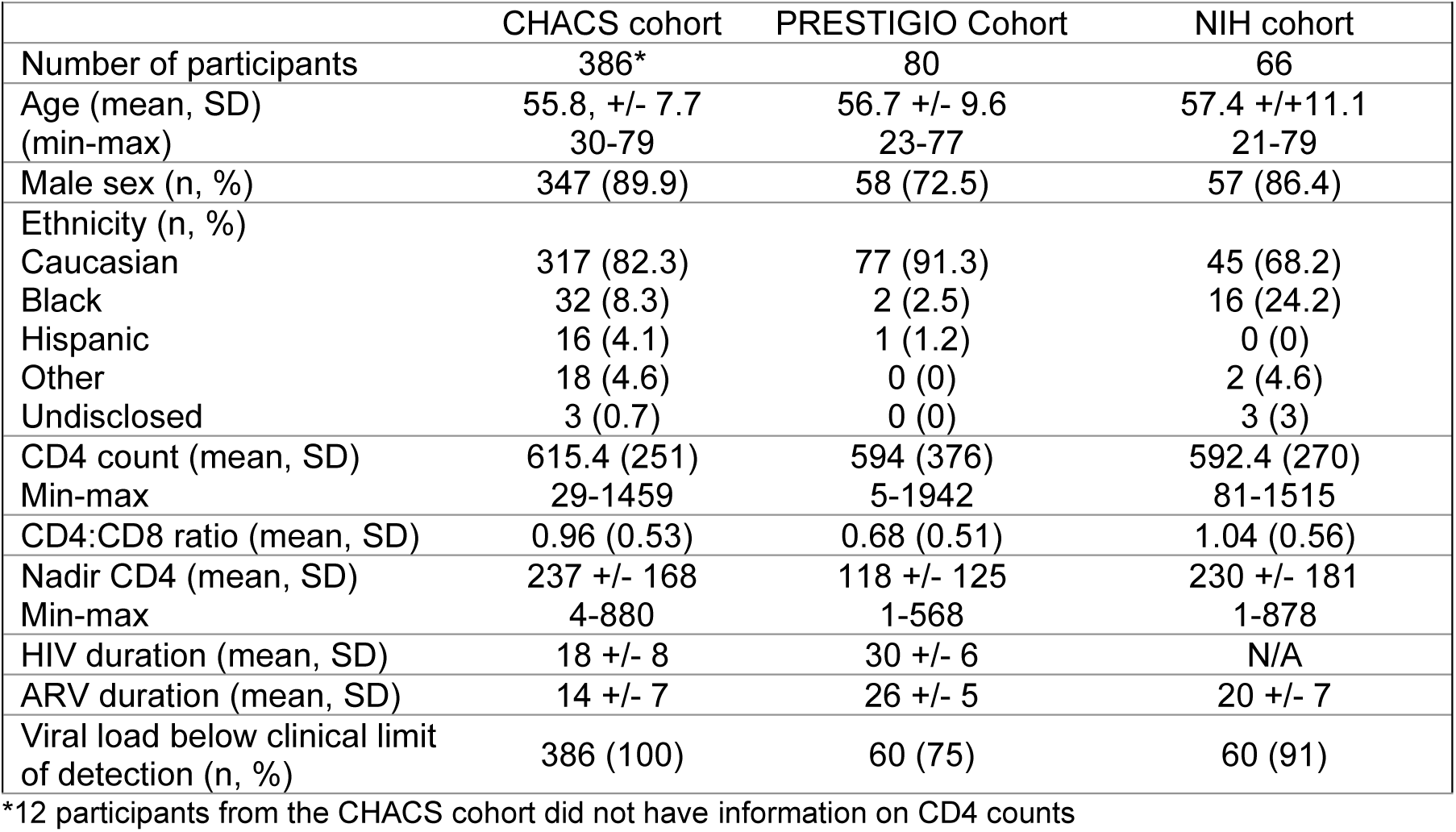
Participants characteristics.

**Supplemental Table 2.**
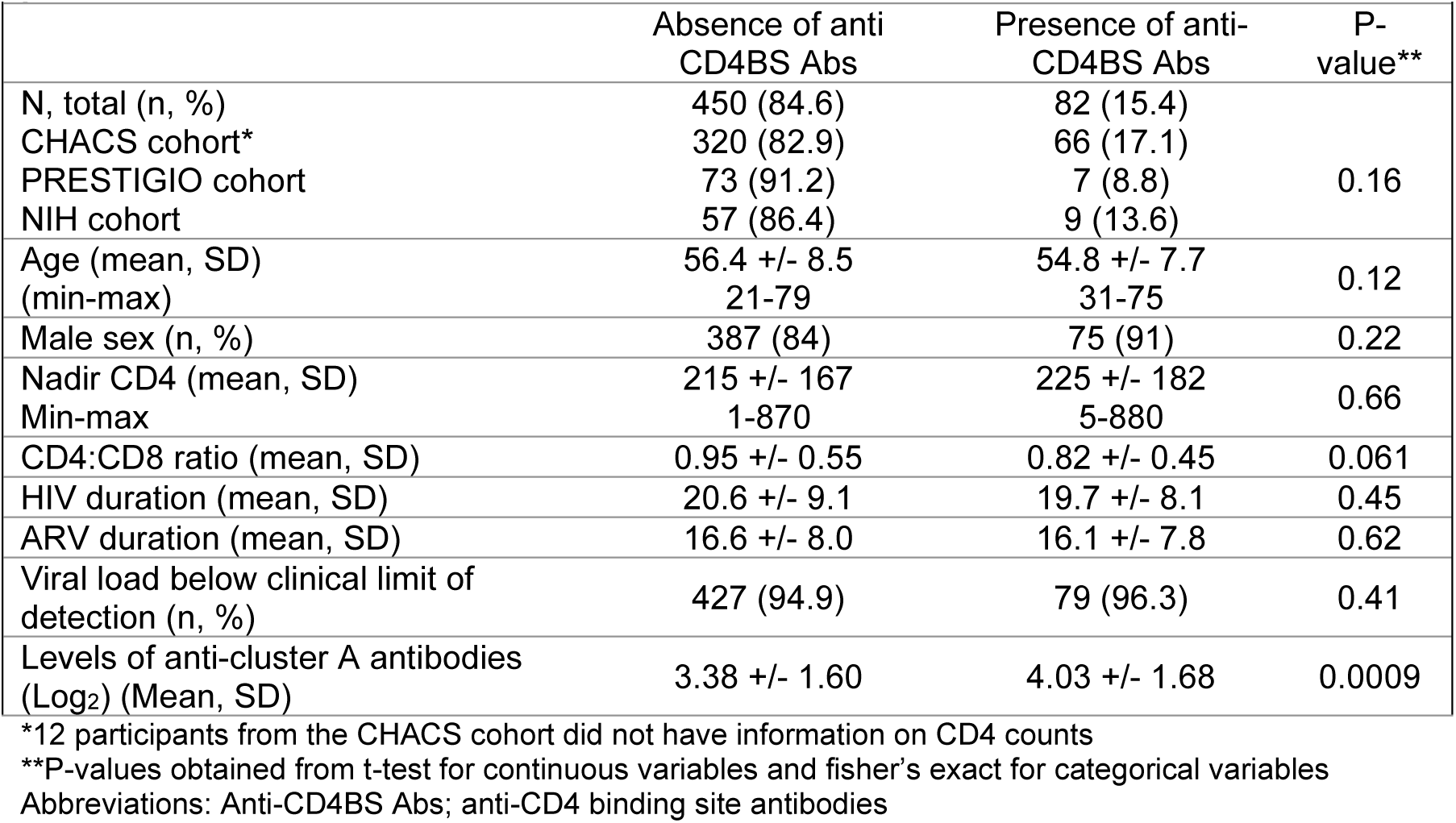
Characteristics of participants stratified by the absence or presence of anti-CD4BS Abs.

**Supplemental Table 3.** Crude associations between anti-cluster A Abs and CD4 count by study cohorts, overall and stratified by the absence or presence of anti-CD4BS Abs.

|  |  |  |  |  |
| --- | --- | --- | --- | --- |
|  | n | Beta*, 95% CI | P-value | P-value for interaction |
| CHACS cohort |  |  |  |  |
| Overall | 374** | -12.04 [-27.81 to 3.74] | 0.13 | N/A |
| Absence of anti-CD4BS Abs | 311 | -16.67 [-34.00 to 0.68] | 0.06 | 0.26 |
| Presence of anti-CD4BS Abs | 63 | 6.77 [-34.45 to 47.99] | 0.74 |  |
| PRESTIGIO cohort |  |  |  |  |
| Overall | 80 | - 47.65 [-98,55 to 3.23] | 0.07 | N/A |
| Absence of anti-CD4BS Abs | 73 | - 68.97 [-120.26 to -17.68] | 0.009 | 0.01 |
| Presence of anti-CD4BS Abs | 7 | 161 [-98.37 to 421.77] | 0.17 |  |
| NIH cohort |  |  |  |  |
| Overall | 66 | -20.09 [-60.57 to 20.39] | 0.32 | N/A |
| Absence of anti-CD4BS Abs | 57 | -27.24 [-70.56 to 21.62] | 0.21 | 0.81 |
| Presence of anti-CD4BS Abs | 9 | - 9.97 [-190.95 to 171.02] | 0.90 |  |

\*beta coefficients represent the predicted change in CD4 count for each increase of 1-log<sub>2</sub> in levels of anti-cluster A antibodies
\*\*12 participants did not have information on CD4 counts and were excluded from the analysis
Abbreviations: CI; confidence intervals, anti-CD4BS Abs; anti-CD4 binding site antibodies
Also refer to Figure 2.

**Supplemental Table 4.** Adjusted associations between anti-cluster A Abs and CD4 count by study cohorts, overall and stratified by the absence or presence of anti-CD4BS Abs.

Also refer to Figure 2.
|  |  |  |  |  |
| --- | --- | --- | --- | --- |
|  | n | Beta*, 95% CI | P-value | P-value for interaction |
| CHACS cohort |  |  |  |  |
| Overall | 374** | -18.17 [-33.34 to -3.01] | 0.019 | N/A |
| Absence of anti-CD4BS Abs | 311 | -23.23 [-39.88 to -0.15] | 0.006 | 0.23 |
| Presence of anti-CD4BS Abs | 63 | 0.08 [-41.07 to 41.24] | 0.99 |  |
| PRESTIGIO cohort |  |  |  |  |
| Overall | 80 | - 36.39 [-88.68 to 15.90] | 0.17 | N/A |
| Absence of anti-CD4BS Abs | 73 | - 61.11 [-114.6 to -7.60] | 0.026 | 0.02 |
| Presence of anti-CD4BS Abs | 7 | 161.69 [-126.74 to 450.13] | 0.19 |  |
| NIH cohort |  |  |  |  |
| Overall | 66 | -28.11 [-66.97 to 10.75] | 0.15 | N/A |
| Absence of anti-CD4BS Abs | 57 | -25.23 [-65.93 to 15.47] | 0.22 | 0.52 |
| Presence of anti-CD4BS Abs | 9 | - 9.97 [-200.54 to 180.59] | 0.90 |  |

\*beta coefficients represent the predicted change in CD4 count for each increase of 1-log<sub>2</sub> in levels of anti-cluster A antibodies
\*\*12 participants did not have information on CD4 counts and were excluded from the analysis
All models are adjusted for age, sex, duration of antiretroviral therapy and nadir CD4
Abbreviations: CI; confidence intervals, anti-CD4BS Abs; anti-CD4 binding site antibodies

**Supplemental Table 5.** Pooled crude associations between anti-cluster A Abs and CD4 count, overall and stratified by the absence or presence of anti-CD4BS Abs.

|  |  |  |  |  |
| --- | --- | --- | --- | --- |
|  | n | Beta*, 95% CI | P-value | p-value for interaction |
| Overall** | 520 | -18.32 [-32.85 to -3.79] | 0.013 | NA |
| Absence of anti-CD4BS Abs | 441 | -26.74 [-42.52 to -10.96] | 0.001 | 0.02 |
| Presence of anti-CD4BS Abs | 79 | 19.77 [-19.42 to 58,95] | 0.32 |  |

\*beta coefficients represent the predicted change in CD4 count for each increase of 1-log<sub>2</sub> in levels of anti-cluster A antibodies
\*\*12 participants did not have information on CD4 counts and were excluded from the analysis
Abbreviations: CI; confidence intervals, anti-CD4BS Abs; anti-CD4 binding site antibodies
Also refer to Table 1.

